# Amygdala connectivity as a predisposing neural feature of stress-induced behaviour during the COVID-2019 outbreak in Hubei

**DOI:** 10.1101/2021.07.26.21261160

**Authors:** Yuan Zhou, Yuwen He, Yuening Jin, Bei Rong, Peter Zeidman, Huan Huang, Yuan Feng, Jian Cui, Shudong Zhang, Yun Wang, Gang Wang, Yutao Xiang, Huiling Wang

**Author notes:** **Corresponding author:** Yuan Zhou, Ph.D., CAS Key Laboratory of Behavioral Science and Magnetic Resonance Imaging Research Center, Institute of Psychology, Chinese Academy of Sciences,16 Lincui Road, Chaoyang District, Beijing 100101, P.R. China; Huiling Wang, M.D., Department of Psychiatry, Renmin Hospital of Wuhan University, 238 Jiefang Road, Wuhan 430060, China.

## Abstract

The amygdala plays an important role in the regulation of stress and anxiety. However, little is known about the relationship between amygdala connectivity and subsequent stress-induced behavior. The current study investigated whether amygdala connectivity measured before experiencing stress is a predisposing neural feature of subsequent stress-induced behavior while individuals face an emergent and unexpected event like the COVID-19 outbreak. Using an fMRI cohort established before the pandemic in Wuhan, Hubei, we found that resting-state functional connectivity (rsFC) of the right amygdala with the dorsomedial prefrontal cortex (dmPFC) was negatively correlated with the stress-induced behavior of these volunteers during the COVID-2019 outbreak in Hubei. Furthermore, the self-connection of the right amygdala, inferred using dynamic causal modeling, was negatively correlated with stress-induced behavior in this cohort. A significant correlation between the right amygdala-dmPFC rsFC and self-connection of the right amygdala was found. Additionally, after three months of the COVID-19 outbreak in Hubei when the stressor weakened - and in another cohort collected in regions outside Hubei where the individuals experienced a lower level of stress - the relationship between the amygdala-dmPFC rsFC and the stress-induced behavior disappeared. Our findings support that amygdala connectivity is a predisposing neural feature of stress-induced behavior in the COVID-19 outbreak in Hubei, suggesting the amygdala connectivity before stress predicts subsequent behavior while facing an emergent and unexpected event. And thus our findings provide an avenue for identifying individuals vulnerable to stress using intrinsic brain function before stress as an indicator.

## 1 Introduction

The novel coronavirus pneumonia (COVID-19) pandemic is affecting people around the world. The COVID-19 pandemic is an uncertain and unpredictable psychosocial stressor (Gruber et al. 2021), which influences both individuals’ physical and mental health (Holmes et al. 2020, Xiang et al. 2020). The increased anxiety, depression, and other negative emotions have been observed not only among the infected patients but also among the general public (Fegert et al. 2020, Vindegaard and Eriksen Benros 2020). Individual differences in reactions to this pandemic-related stressor has also been observed by some studies (Di Crosta et al. 2020, Zacher and Rudolph 2021), which are similar to the previous observations that some individuals are more vulnerable to stressors or traumatic events than others (Breslau et al. 1991, Bolsinger et al. 2018). In this context, a better understanding of the neural basis of stress-induced behavior could facilitate precise identification of the vulnerable individuals and develop preventive interventions on these individuals.

Previous animal and human studies have demonstrated the importance of the amygdala in stress responses and stress disorders (e.g. post-traumatic disorders, major depressive disorders, generalized and social anxiety disorders, etc.), due to its role in the regulation of fear and anxiety (Etkin and Wager 2007, Chattarji et al. 2015; Rauch et al. 2000, Stein et al. 2007, Shin and Liberzon 2010). A rising concern in this field of study is the direction of influence, namely whether amygdala activity is a neurobiological vulnerability marker for stress-related disorders or a consequence of stress exposure / stress-related disorders (for a review, please see also Disner et al. 2018). For both directions of influence there are a few supportive evidence. Studies that supported the stress-to-amygdala direction of influence primarily investigated the role of early life stressors on abnormal amygdala connectivity in later lives (McLaughlin et al. 2019, Herzberg and Gunnar 2020), how experimental manipulation of acute stress influences amygdala activities (Chang and Yu 2018, Orem et al. 2019), and how stress-related psychiatric disorders alter amygdala functions (Shin et al. 2005, Kim et al. 2011, Robinson et al. 2014, Hultman et al. 2016, Zhang et al. 2016). On the other hand, supportive evidence on the amydala-to-stress direction of influence is scarce (Admon et al. 2009, McLaughlin et al. 2014), due to two major difficulties in experimental design. First, the time of occurrence of natural stressors cannot be foreseen. Second, the acquisition of amygdala connectivity data before the occurrence of natural stressors is difficult. Although several experimental studies have attempted to overcome the difficulties by investigating how task-induced amygdala’s activity led to vulnerability to trauma exposure (Admon et al. 2009, McLaughlin et al. 2014). However, they cannot capture intrinsic individual differences in amygdala connectivity or responses to real chronic stressors in natural settings. Therefore, it is still unknown whether amygdala connectivity long before experiencing stress is a predisposing neural feature of subsequent stress-induced behavior while facing an emergent and unexpected event, like the COVID-19 outbreak.

Resting-state fMRI is a powerful tool to uncover individual differences in human cognitive abilities and behavioural tendencies (Raichle et al. 2001, Raichle 2010). Spontaneous activity during rest in functionally related brain regions, measured by resting-state functional connectivity (rsFC), is intrinsically organized, revealing the functional architectures of the brain (Fox and Raichle 2007, Smith et al. 2009). The spontaneous brain activity has great consistency across time in previous studies, and has proved to be a potential predictor of individual differences in behavioral tendencies (Harmelech and Malach 2013, Finn et al. 2015, Tavor et al. 2016). Two studies from one group have found that the rsFC can predict feelings of stress or anxiety related to the COVID-19 pandemic (He et al. 2021, Liu et al. 2021); however, neither of the studies focused on the amygdala connectivity.

Animal studies have demonstrated that the directed interaction between amygdala and other brain regions, such as dorsomedial prefrontal cortex (dmPFC), is highly correlated with the increased anxiety-like behavior in stressed mice (Liu et al. 2020). To capture how the directional interaction between amygdala and brain regions predict subsequent stress-induced behavior in humans, we incorporated the dynamic causal modeling (DCM), a widely adopted framework for effective connectivity analysis (Friston et al. 2003). DCM can better disclose the causal and directed nature of coupling between intrinsic modes of brain activity, as a nice complementary to rsFC, which only describes a non-directed functional interaction between fMRI time series across the brain and does not reveal the causal influence of one neural system (Friston 2011). Technologies called *stochastic* and *spectral* dynamic causal modeling (spDCM) are the most recent approaches to characterize effective connectivity during rest (Friston et al. 2014). Compared to its stochastic counterpart, spDCM, which operates in the frequency domain rather than the time domain, is more computationally efficient and more accurate and sensitive to group differences (Razi et al. 2015). Therefore, we used the spDCM to estimate effective connectivity that underlie intrinsic functional connectivity during rest (Razi and Friston 2016) and test the hypothesis that the directed connectivity of amygdala can predict subsequent stress-induced behavior as a predisposing neural feature.

This study grasps the unique chance of COVID-19 outbreak and leverages data from the established cohort to investigate whether resting-state functional and effective connectivity of amygdala measured before the COVID-19 pandemic is a predisposing neural feature for subsequent stress-induced behavior in the individuals in the geographic pandemic center (i.e. Hubei Cohort), who were exposed to high level of pandemic stressors. This study further investigates whether such predictors of response still could be used to infer the stress-induced behaviors when the stressor weakened (i.e., after three months of the COVID-2019 outbreak in Hubei) and whether such predictors of response exist among another group (non-Hubei Corhort) in which individuals were exposed to a low level of pandemic stressors.

## 2 Materials and methods

### 2.1 Participants^1^

The main analyses of this study were conducted based on the Hubei Cohort, which consists of the normal controls in a previous fMRI study of schizophrenia which was conducted at the Renming Hospital, Wuhan University from June, 2012 to July, 2019. The data have been used in previous studies (Huang et al. 2018, Li et al. 2020). In these neurologically normal participants, 50 participants took part in the current study. Among of them, 45 participants lived in Hubei province when they were recruited in the original project, and they were living or still lived in Hubei province half a year before the COVID-19 outbreak in Hubei - the pandemic center of the COVID-19 in China - at the time of questionnaire data collection of this study (Feb 15th and 29th, 2020). Thus, these participants were assumed to experience a high level of pandemic stressors.

To test whether participants in the pandemic center display higher stress-induced behavior than those not in the pandemic center, we recruited fifty-eight volunteers from another established non-clinical pool of a study conducted in Beijing (non-Hubei Cohort). All of these participants resided outside the Hubei province when they were recruited in the original project and half a year before COVID-19 outbreak in Hubei. Because there were much fewer COVID-19 cases in other provinces outside Hubei province at the time of questionnaire data collection of this study (from Feb 15th to 29th, 2020) (http://www.nhc.gov.cn/xcs/yqtb/list_gzbd.shtml), this cohort was assumed to experience a low level of pandemic stressors.

All participants had reported no history of major psychiatric or neurological illness when they were recruited. All of the participants were invited to complete an online survey questionnaire via text messages during Feb 15th and 29th, 2020. Importantly, based on their self-reports, none of participants in the cohort were suspected cases or patients with the COVID-19. This study was approved by the Ethics Committee of Renmin Hospital of Wuhan University, and the Institutional Review Board of the Institute of Psychology, Chinese Academy of Sciences. All of the subjects gave informed consent online.

### 2.2 Measurements on COVID-19 related stress behaviors

A 14-item self-report questionnaire, Stress Behavior Scale induced by COVID-19 (SBSC), was developed to assess COVID-related stress behaviors following self-report scale development principle by Tay and Jebb (Tay and Jebb 2017). The detailed procedures were included in supplementary materials.

We also collected the self-report measurements on anxiety, stress and depression during this survey using existed scales, including a Chinese version of the State-Trait Anxiety Inventory (S-TAI) (Spielberger 1989), the Perceived Stress Scale-10 (PSS-10) (Cohen 1988) and the Patient Health Questionnaire-9 (PHQ-9) (Kroenke et al. 2001).

### 2.3 MRI data acquisition and analyses

#### 2.3.1 Imaging protocol

MRI scanning was performed on a 3.0T General Electric Signa HDxt MR scanner in the Department of Radiology, Renmin Hospital of Wuhan University. Resting-state functional images were obtained by using an echo-planar imaging (EPI) sequence (repetition time [TR] = 2000 ms, echo time [TE] = 30 ms, flip angle = 90°, field of view [FOV] = 220 mm × 220 mm, matrix = 64 × 64, 32 slices, slice thickness = 4 mm, and gap = 0.6 mm) and 240 volumes were obtained. Structural images were collected using a 3D Bravo T1-weighted sequence (TE=7.8 ms, TR = 3.0 ms, flip angle = 7°, inversion time = 1100 ms, FOV = 256 mm × 256 mm, matrix = 256 × 256, 188 slices, and voxel size 1 mm × 1 mm × 1 mm). During the resting-state scanning for both of the cohorts, all participants were instructed to close their eyes and to focus on nothing in particular. For details of the imaging protocol in the non-Hubei group, please see the supplementary materials.

#### 2.3.2 Preprocessing

All imaging data preprocessing procedures were carried out with Data Processing Assistant for Resting-state fMRI version 4.3 (http://www.restfmri.net), which is based on Statistical Parametric Mapping 12 (http://www.fil.ion.ucl.ac.uk/spm). We performed the following preprocessing steps: removing the first 5 time points, slice-time correction, realignment, co-registration, segmentation for structural images, nuisance covariates regression (including Friston’s 24 parameters of head motion, 5 principal components from the individual segmented white matter, cerebrospinal fluid, linear and quadratic trends), normalization to MNI space (2 × 2 × 2 mm^3^ cubic voxels) and spatial smoothing with a 4mm FWHM kernel. For resting-state functional connectivity, a temporal filtering (0.01-0.1 Hz) was conducted. For effective connectivity, the voxels showing low frequency fluctuations were identified using a general linear model (GLM) containing a discrete cosine basis set with frequencies ranging from 0.0078 to 0.1 Hz based on previous studies (Razi et al. 2015, Almgren et al. 2020). An F-contrast was specified across the discrete cosine transforms (DCT), producing an SPM that identified regions exhibiting BOLD fluctuations within the frequency band. A grey matter mask was generated by including the voxels in which 90% of subjects contained EPI signal and the mean gray matter values were larger than 0.2.

Because of the potential influences of head motion on the rsFC, we further reduced the influences of motion-related artifacts on rsFC. First, we used volume-based framewise displacement (FD) to quantify head motion (Power et al. 2012). The time points with a threshold of FD > 0.5mm as well as 1 back and 2 forward frames were identified as “bad” points and then were modeled as a separate regressor in the regression model of the realigned resting fMRI data while removing nuisance covariates (Yan et al. 2013). Second, we excluded participants who had less than 120 “good” volumes of data (Yan et al. 2013) or whose mean FD was above 3 standard deviations beyond the mean value of the whole sample. No participants were excluded. Finally, the mean FD was entered into the group analyses as one of nuisance covariates.

#### 2.3.3 Functional connectivity analyses

The left and right amygdala derived from the SPM Anatomy toolbox (Eickhoff et al. 2005, Eickhoff et al. 2006) were used as the seed regions for seed-based rsFC analysis separately. We calculated the Pearson correlation between the mean time series of the seed and the time series of each voxel within the grey matter mask. After transforming the Pearson correlations into z-values, the resulting z-valued functional connectivity maps of each seed were entered to the multiple regression analyses to investigate the relationship between amygdala’s functional connectivity and the SBSC score. To remove the confounding effects, we included gender, age and mean FD as covariates to the regression model. Statistical significance was set at a cluster-defined threshold *p* < 0.001 in conjunction with cluster wise FWE *p* < 0.025 (Bonferroni correction for two seed-based rsFC analyses) to correct for multiple comparisons.

#### 2.3.4 Effective connectivity analyses

According to the results of functional connectivity analyses, spDCM was used to reveal the relationship between directed connectivity and stress behaviors induced by the COVID-19 pandemic. Specifically, we took the right amygdala and the dmPFC as the volume of interest (VOI). The principal eigenvariate of the voxels in each VOI was computed separately. Then a spDCM analysis was conducted using DCM12.5 implemented in the SPM12 (revision 7497, www.fil.ion.ucl.ac.uk/spm). For each participant, a fully connected model was created to answer the question of whether the stress-induced behavior was influenced by the top-down regulation effect from the dmPFC to the right amygdala, or the down-up regulation effect from the right amygdala to the dmPFC. We were also interested in the self-connections within each region, which can be considered as parametrizing the interplay between inhibitory interneurons and pyramidal cells within a brain region, thus reflecting the gain or excitability of neuronal populations reporting prediction errors (Clark 2013, Friston et al. 2014, Ranlund et al. 2016). Therefore, we constructed a full model consisting of the directed connections between the right amygdala and the dmPFC as well as self-connections within each region.

The inversion of DCM at the first level was performed using spDCM, which fits the complex cross-spectral density using a power-law model of endogenous neuronal fluctuations (Friston et al. 2014, Razi et al. 2015). Then we used Parametric Empirical Bayes (PEB) (Zeidman et al. 2019) to model how individual connections relate to group means and individual differences in stress-induced behavior related to COVID-19 outbreak indicated by the SBSC scores. After inverting the PEB model (spm_dcm_peb.m), we used Bayesian Model Reduction to find out whether there was an effect of SBSC scores on the effective connectivity and, if so, where it was expressed (Friston et al. 2016, Zeidman et al. 2019). In the PEB framework, group-level analysis is conducted using Bayesian inference. It avoids the need to contend with the multiple-comparison problem of classical inference, because the objective is to quantify the posterior probability for effects, rather than determine whether they exceed a significance threshold (Friston and Penny 2003).

We computed the Bayesian posterior probability for our effects of interest using variational Bayesian methods, as implemented in the PEB framework. Then, we computed the Bayesian Model Average, which is the average of the parameters from different models weighted by the models’ posterior probabilities, to present the results. Here we focus on effects with posterior probability >0.95, which is considered “strong evidence” for an effect (Kass and Raftery 1995).

### 2.4 Follow-up analyses

We conducted the second survey three months after the first survey, when the COVID-19 pandemic had been effectively controlled in China, as indicated by the fact that Wuhan, a Hubei city which was the most seriously affected by the COVID-19, lifted lockdown on 8 April, 2020 due to the sharp reduction of daily increased diagnosed cases. During the second survey (from May 28 to June 8, 2020), we collected the data from 33 participants in the Hubei Cohort and 58 participants in the non-Hubei Cohort using the same questionnaires as the first survey. We repeated the abovementioned analyses to explore the correlations between the amygdala connectivity and the SBSC scores.

### 2.5 Complementary analyses

While the primary aim of including the non-Hubei Cohort was to validate the SBSC questionnaire, we also applied the same functional connectivity analyses to their fMRI data, to test whether the relationship between amygdala connectivity and stress-induced behavior exists among individuals who were exposed to low level of pandemic stressors (for more details, please see also supplementary materials). However, as the two cohorts were unavoidably scanned using MRI scanners in different locations (Hubei and Beijing), we could not perform a formal comparison of the rsFC across cohorts. We therefore present their results separately (section 3.5).

## 3 Results

### 3.1 Measurements on COVID-19 related stress behaviors and emotions

There was no significant difference in the gender composition and education level, while a significant difference in age, existed between the Hubei Cohort and non-Hubei Cohort (*p* < 0.05) (Table 1). Regression analyses after controlling for age showed that participants in the Hubei Cohort had significantly higher scores in the COVID-19 related stress behaviors and several measures on anxiety and perceived stress than the non-Hubei Cohort. There was no significant difference in the level of depression between the two groups (Table 1).

**Table 1.** Demographic and behavioral comparisons between the research group and the control group

|  | Research Group<br>(*N*=45)<br>(*Mean (SD)*) | Control Group<br>(*N*=58)<br>(*Mean (SD)*) | Group Difference Statistics |
| --- | --- | --- | --- |
| Age (year) | 29.38 (4.10) | 25.85 (3.53) | $t(101)=4.69, p < .001$ |
| Gender (male/female) | 22/23 | 17/41 | $\text{Chi-square} = 4.13, p = .04$ |
| Education (year) | 16.767 (4.19) | 17.97 (2.60) | $t(101)=-1.82, p=.07$ |
| Head motion (mean FD) | 0.12 (0.06) | 0.13(0.007) | $t(101)=-1.28, p=0.20$ |
| PHQ-9 | 4.16 (3.81) | 3.26 (4.37) | $t(101)=1.09, p=.28$ |
| TAI | 40.93 (7.69) | 36.17 (9.96) | $t(101)=2.74, p=.01$ |
| SAI | 37.58 (11.41) | 31.98 (11.11) | $t(101)=2.50, p=.01$ |
| PSS | 15.84 (4.58) | 12.02 (6.17) | $t(101)=3.48, p=.001$ |
| COVID-19 related stress behaviors | 34.73 (13.44) | 22.93 (9.76) | $t(101)=4.96, p<.001$ |

Note: There is significant difference between the research and the control group on mean age level. We compared the level of PHQ-9, TAI, SAI, PSS and COVID-19 related stress-like behavior between the two groups after controlling for age. Abbreviations: PHQ-9, Patient Health Questionnaire-9; PSS, Perceived Stress Scale; NA, no applicable; SAI, State Anxiety Inventory; TAI, Trait Anxiety Inventory.

### 3.2 Functional connectivity

We found the connectivity between the right amygdala and the dmPFC (peak coordinates: [8,48,36], cluster size = 172 voxels, cluster-level FWE *p* = 0.001) was negatively correlated with the SBSC scores, suggesting that the individuals with weaker correlation between the right amygdala and the dmPFC exhibited more stress-induced behavior (Figure 1). No correlations between the rsFC of the left amygdala and the SBSC scores were found (cluster-level FWE *p* < .025).

**Figure 1.**
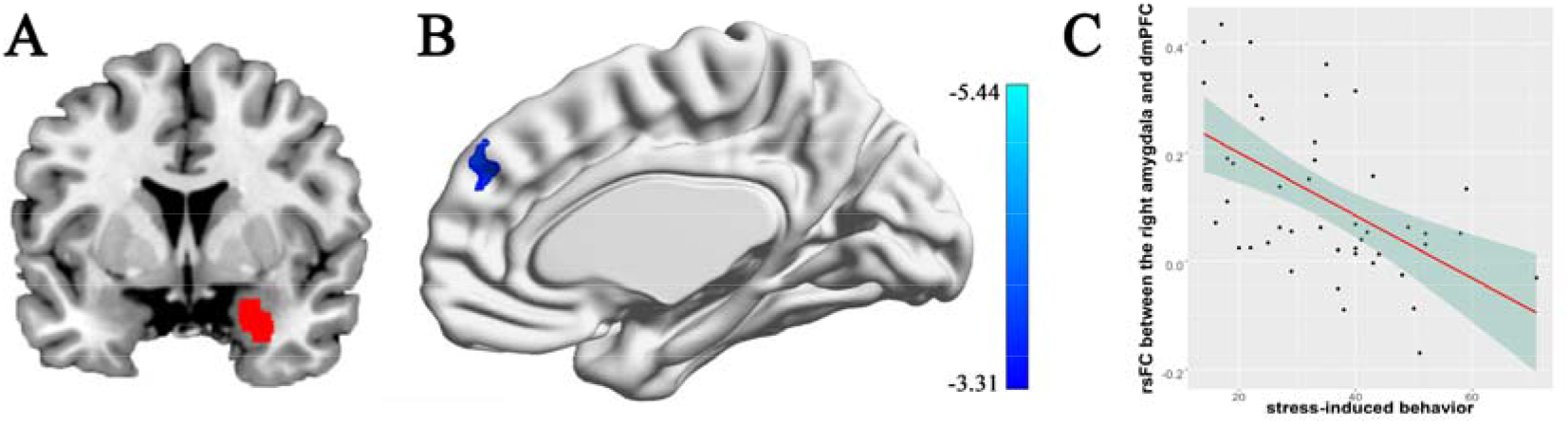
Correlation between rsFC of the right amygdala in the dmPFC and stress-induced behaviors. (A) The right amygdala; (B) The dmPFC whose rsFC with the right amygdala correlated with the stress behaviors during the COVID-19 outbreak; (C) A scatter plot showing the relationship between the rsFC and the stress-induced behaviors.

### 3.3 Effective connectivity

We found that in common across participants, as illustrated in Figure 2A, there was inhibitory connectivity from the right amygdala to the dmPFC and excitatory connectivity from the dmPFC to the right amygdala, which exceeded 95% posterior probability of being present v.s. absent, based on comparing the approximate log evidence for models with and without each connectivity parameter. Also, the level of self-inhibition was greater than was expected under the priors for both regions, as shown by the positive parameter estimates. More importantly, we found a negative effect of the SBSC scores on the inhibitory self-connection of the right amygdala (posterior probability > 95%), showing that individuals with weaker self-inhibition of the right amygdala had more stress-induced behaviors (Figure 2B).

**Figure 2.**
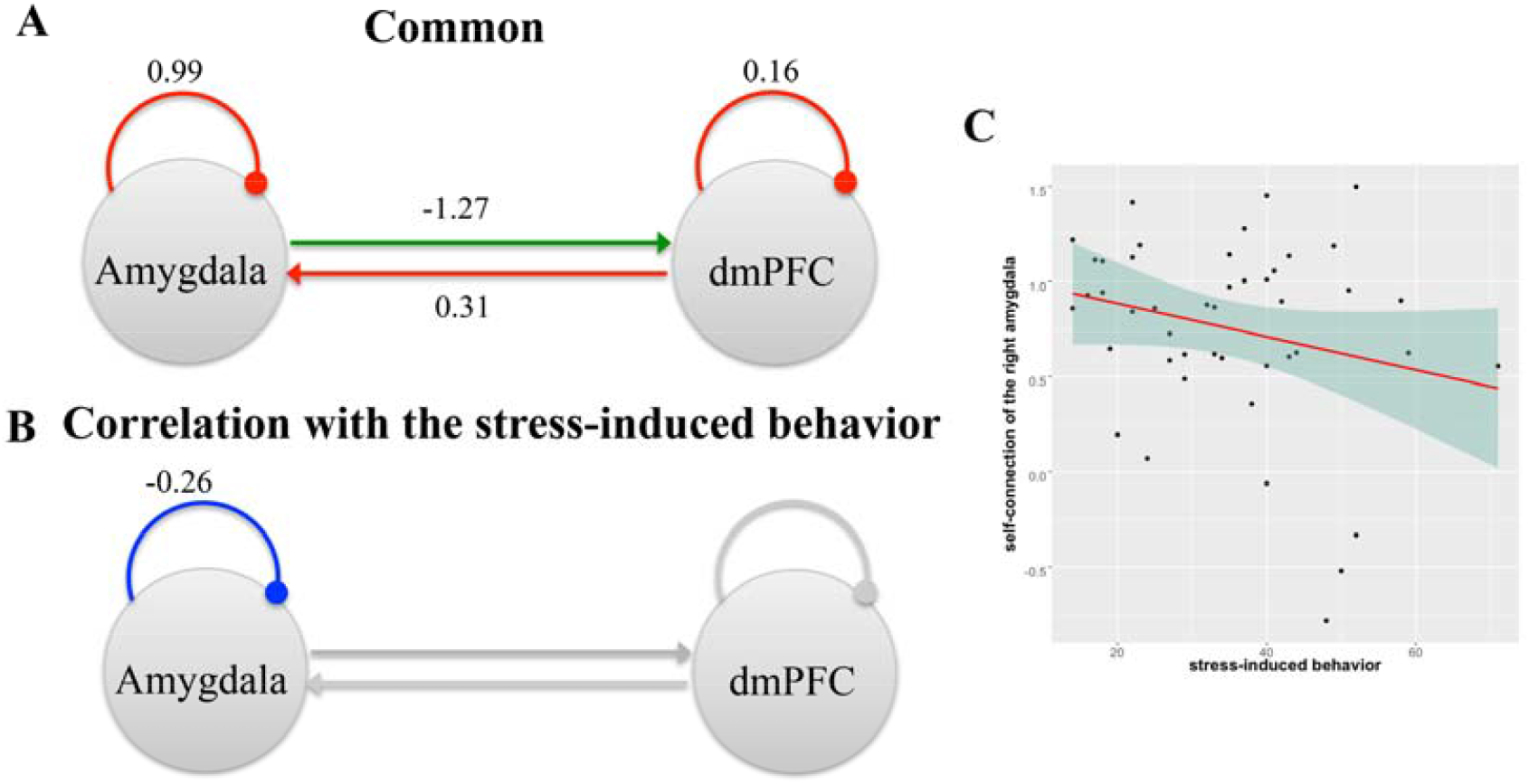
A schematic summarizing effective connectivity between the right amygdala and the dmPFC across participants in the Hubei Cohort and its correlation with stress-induced behavior. (A) The green arrow represents the positive extrinsic effective connectivity and the red arrow represents the negative extrinsic effective connectivity. The parameters are the strength of connectivity. The red arcs represent self-connections. For the self-connections, the parameters are log scaling parameters, which can be converted to units of Hz by: y =**−**0.5 * exp(x). Where x is the log scaling parameter, **−**0.5 Hz is the prior and y is the self-connection strength in units of Hz. (B) The parameter is the effect of the stress behaviors on the self-connection. All of the other connections in this network, which had no correlations with the behaviour, were shown in grey. (C) A scatter plot showing the relationship between the self-connection in the right amygdala and the stress-induced behaviors.

To aid in interpreting the relationship between effective connectivity and functional connectivity, an analysis was conducted to examine whether the self-connection of the right amygdala was associated with the amygdala-dmPFC functional connectivity. We found that individuals with weaker self-connection of the right amygdala had less amygdala-dmPFC functional connectivity (r-0.38, p=0.01) (Figure 3), possibly due to impaired self-inhibition of amygdala (disinhibition or hyper-activity).

**Figure 3.**
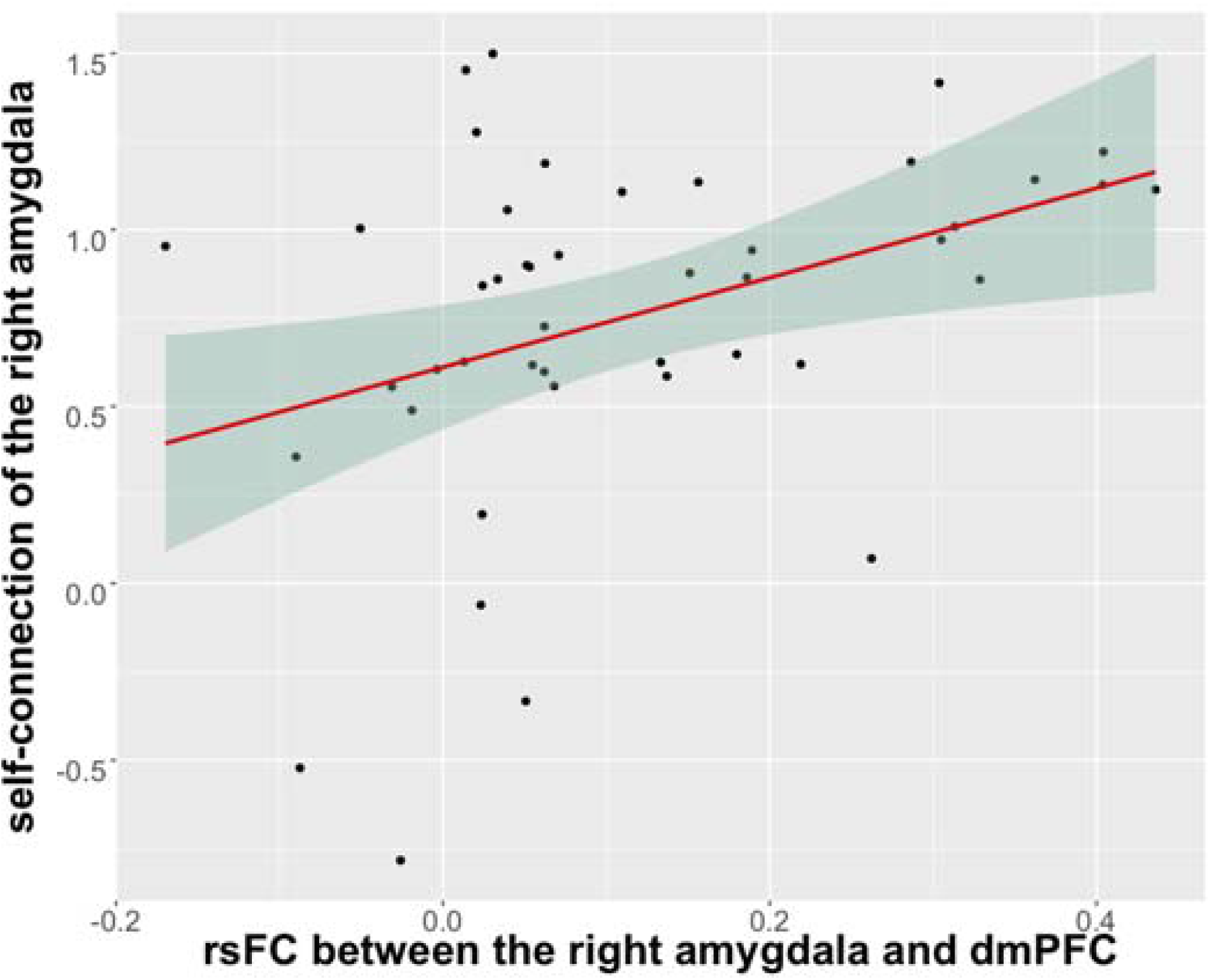
A scatter plot showing the relationship between the self-connection in the right amygdala and the rsFC of the right amygdala in the dmPFC.

### 3.4 Follow-up analyses

After three months, when the COVID-19 pandemic had been effectively controlled in China, stress-induced behaviors measured by the SBSC scores decreased in the Hubei Cohort (25.61±10.80) relative to the first survey (31.94±12.34) in the 33 participants who completed the second survey, but the score is still higher than that in the non-Hubei Cohort (16.95±5.04; t = 4.34; *p* < 0.001). In addition, significant time effects (F=34.53, p<0.001) were revealed in these two groups, while the interaction between time and site was not significant (F=0.02, p=0.89) after a repeated measure ANOVA analysis.

Using the data of these 33 participants in the Hubei Cohort, we could still find a negative correlation between the connectivity of the right amygdala in the dmPFC and the SBSC scores of the first survey (*p* = 0.001, cluster-wise FWE *p* < 0.025, Figure S1 in the supplementary materials), which validated our main finding in a smaller sample size. There is no significant correlations were found between the amygdala’s rsFC and the SBSC scores of the second survey in the Hubei Cohort in the dmPFC. Instead, it was found that a cluster in the superior frontal gyrus (SFG) of the right amygdala’s rsFC was negatively correlated with SBSC (*p* = 0.001, cluster-wise FWE *p* < 0.025, Figure S2 in the supplementary materials).

### 3.5 Complementary analyses

No correlations between the rsFC of the left or right amygdala and the SBSC scores were found in non-Hubei Cohort (*p* = 0.001, cluster-level FWE *p* < 0.025). In addition, no significant correlations were found between the amygdala’s rsFC and the SBSC scores in the second survey in the non-Hubei Cohort (cluster-level FWE *p* > 0.05). Of course, failure to reject the null hypothesis does not provide evidence for the null, and it was not possible to formally compare the rsFC across cohorts as they were scanned using different scanners (Beijing and Hubei). Nevertheless, null results in the non-Hubei group are what one would expect to see, given the strong association between amygdala rsFC and SBSC scores in the Hubei group.

## 4 Discussions

This study demonstrated that amygdala connectivity during rest obtained before the COVID-19 pandemic can be taken as a predisposing neural feature for stress-induced behavior during the COVID-19 outbreak in Hubei. Specifically, in an existing cohort of non-clinical population, we found that the functional connectivity of the right amygdala in the dmPFC was negatively correlated with the scores of stress-induced behavior; guided by this finding, we further found that the self-connection of the right amygdala was correlated with the scores of stress-induced behavior, suggesting that the individuals with a weaker self-inhibition (i.e., disinhibition, or hyper-activity) of the right amygdala before the pandemic are at a greater risk to exhibit stress-induced behaviors during the COVID-19 outbreak.

Previous studies have reported hyperactivity within the amygdala and impaired functional connectivity between the amygdala and mPFC in stress-related psychiatric disorders (Shin et al. 2005, Kim et al. 2011, Robinson et al. 2014, Hultman et al. 2016) and found that early stress can alter the amygdala’s connectivity (McLaughlin et al. 2019, Herzberg and Gunnar 2020). Two studies heightened that amygdala reactivity to tasks is related to vulnerability to trauma-related psychopathology (Admon et al. 2009, McLaughlin et al. 2014). The current study expanded our understanding on the relationship between amygdala and stress in two important ways. First, taking advantage of an existing cohort built before the COVID-19 pandemic, we have the chance to find the evidence that the spontaneous brain activity of amygdala before stress can predict the subsequent stress-induced behaviors while facing a public health emergency, suggesting that amygdala function is a neural vulnerability factor to a stressful event exposure in addition to being the consequence of a pathological response to the exposure. Secondly, using effective connectivity, we provide the first evidence for the role of the self-connection in the amygdala in predicting subsequent stress-induced behaviors.

The functional connectivity between the amygdala and the dmPFC has been recognized in stress-related emotions and stress-related psychiatric disorders (Quirk et al. 2003, Kim et al. 2011, Robinson et al. 2014, Hultman et al. 2016, Liu et al. 2020). As a part of mPFC, which is the primary brain region responsible for processing stressful events or adversity (Arnsten 2009, Liu et al. 2020), the dmPFC is the key neural substrate of the conscious appraisal of threat and subsequent regulation of fear and anxiety (Kalisch and Gerlicher 2014). Convergent evidence from animal and human studies also suggest that the dmPFC is implicated in the expression of threat-related defensive response (Etkin et al. 2011). Furthermore, the dmPFC mediates the impact of serotonin transporter linked polymorphic region genotype on anticipatory threat reactions (Klumpers et al. 2015). More generally, the dmPFC has been associated with outcome evaluation and strategic control under uncertainty (Yoshida and Ishii 2006, Venkatraman and Huettel 2012). The negative correlation between the rsFC between the amygdala and dmPFC and the stress-induced behavior during the COVID-19 outbreak suggests that an individual with weaker rsFC between the amygdala and dmPFC before stress is more likely to have stress-induced behavior during the COVID-19 outbreak. This negative correlation pattern is consistent with a previous study, in which the amygdala—dmPFC coupling during rest is inversely correlated with the avoidance symptom severity of patients with PTSD (Wolf and Herringa 2016). It is generally compatible with the observation of more negative functional coupling between the amygdala and the mPFC when patients with high level of PTSD symptoms were exposed to unpleasant stimuli compared to those with low level of PTSD symptoms (Sadeh et al. 2014). In accordance with this line of studies, our finding, the negative correlation between the amygdala-dmPFC rsFC before stress and the stress-induced behavior during the COVID-19 outbreak, suggests that the brain is ready for future threat or uncertainty by coupling the amygdala and the dmPFC.

Using DCM, we found a common model with an inhibitory connectivity from the right amygdala to the dmPFC and an excitatory connectivity from the dmPFC to the right amygdala. Although functional coupling between amygdala and dmPFC has been repeatedly reported, the directionality of the functional interactions between the two regions has only been examined in few studies. During processing of negative emotion, researchers found that the connectivity from the right amygdala to dmPFC was significant in healthy controls using a method called Granger causality modeling (Lungu et al. 2015, Potvin et al. 2017). A recent study, which also used the spDCM as we did, found inhibitory connectivity from the right amygdala to the dmPFC and the excitatory connectivity from the dmPFC to the right amygdala, as well as the self-connections in both of the two regions, in healthy volunteers during rest in a network including 6 other regions besides the amygdala and dmPFC (Ishida et al. 2020).

We found the self-connection in the right amygdala was related to the stress-induced behavior during the COVID-19 outbreak. In the DCM framework, self-connections are, a priori, constrained to be inhibitory (Friston et al. 2014). This reflects the fact that inhibitory interneurons are restricted to intrinsic anatomical connectivity within the cortex. This means that an increase in self-inhibition corresponds to a reduction in the excitability of neuronal populations to their afferents (and recurrent self-connections) and on the contrary, a decrease in self-connection (i.e. disinhibition) corresponds to an increase in the excitability. Computational accounts under predictive coding interpret these changes in excitability as a failure to attenuate or modulate the precision of prediction errors; namely, the postsynaptic sensitivity of neuronal populations thought to encode prediction errors: e.g., superficial pyramidal cells (Bastos et al. 2012, Shipp 2016). In these accounts, the ensuing psychopathology is often related to an imbalance between sensory and prior precision at lower and higher levels in the cortical hierarchy, respectively. The association with reduced self-inhibition (i.e., disinhibition) and the expression of stress-induced behaviours we observed is particularly interesting in light of predictive coding formulations of stress and anxiety (Cornwell et al. 2017, Peters et al. 2017). The predictive processing formulations of aberrant interceptive inference (Seth and Friston 2016) – in anxiety and stress – often focus on a failure to attenuate the precision of (interoceptive – and related) prediction errors. This results in a hypersensitivity to interoceptive autonomic afferents. Therefore, our findings suggest that individuals with weaker self-inhibition (i.e., disinhibition or hyperexcitability) of the right amygdala would express more stress behaviors, implying the importance of disinhibition / hyperexcitability of the right amygdala in the expression of stress-induced behaviors. Previous studies have already shown that changes in the local regulation of amygdala excitability underlie behavioral disturbances in stress-related psychiatric disorders or stress-induced behaviors (e.g., relapse to drug use) (Prager et al. 2016, Sharp 2017) and chronic stress causes amygdala output neurons to become hyperexcitable (Roozendaal et al. 2009, Rosenkranz et al. 2010, Sharp 2017, Zhang et al. 2019). However, in these previous studies, the hyperexcitability in the amygdala was observed while facing stressors or after experiencing stress. Our current study extends our knowledge on the role of hyperexcitability (i.e., disinhibition) of the amygdala in stress by finding that individuals with hyperexcitability in the amygdala before facing stressors will show more stress-induced behaviors. It is possible that the hyperexcitability in the amygdala makes the individuals more vulnerable to uncertainty and unpredictability of environment and thus more likely to take actions while facing stressors.

It is interesting that we found the strength of self-connection in the right amygdala was positively correlated with the strength of functional connectivity between the right amygdala and the dmPFC. Combining this finding with the inhibitory connectivity from the right amygdala to the dmPFC across participants, we speculate that the hyperexcitability (i.e., disinhibition) in the right amygdala leads to its stronger inhibition effect to the dmPFC, and thus makes the functional coupling between the right amygdala and the dmPFC more towards negative (weaker) connectivity. Because both the weaker self-connection of the right amygdala and the weaker functional connectivity of the right amygdala in the dmPFC were correlated with the more stress behavior, our findings suggest that the role of local excitability-inhibition balance of the amygdala and its connection with the dmPFC in stress are worthy of further investigation in future studies using animal models.

It should be noted that we did not find the relationship between the amygdala-dmPFC connectivity with the stress behavior when the stressor was not so strong, as shown after the stressor weakened, i.e., after three months of the COVID-19 outbreak when Wuhan has lifted lockdown, or in the provinces that had much fewer COVID-19 cases (non-Hubei Cohort). It is easily understood that when the stressor weakens, the responses or behaviors induced by the stressor naturally decrease. Thus these null findings further support that the amygdala connectivity (self-connection and functional connectivity with the dmPFC) is a predisposing neural feature of stress-induced behavior, which is induced by the COVID-19 outbreak in Hubei.

Several limitations should be mentioned. First, the sample size in this study is small because the volunteers were recruited from established cohorts. For the same reason, this study is a quasi-experiment condition and thus we cannot find an appropriate control group to conduct formal statistics to test whether the brain-behavior relationship established in the group with higher stress level (i.e., Hubei Cohort) is statistically stronger than that with lower stress level. An ideal control group could be the individuals who took part in the scanning before the pandemic in the same site as the Hubei Cohort but they or their families were not living in Hubei province half a year before the COVID-19 outbreak in Hubei and thus they were presumed free of the pandemic influence or had lower stress level (there were only 5 cases in the current study). Future studies may use established (e.g., HCP or UK Biobank) or new cohorts with large sample size and diversified sample pool to validate the current findings. Second, this study suggests the role of amygdala connectivity in predicting stress behaviors in a non-clinical population. However, whether the current findings can be generalized to patients with stress-related psychiatric disorders or other vulnerable populations to stress needs to be explored. Furthermore, the physiological mechanism behind this prediction role of amygdala connectivity needs to be explored in future studies by using comprehensive computational model or animal models.

## 5 Conclusions

In conclusion, our findings support the role of functional coupling between the amygdala and dmPFC in stress-induced behaviors and provide new evidence that individuals with hyperexcitability (i.e., disinhibition) in the amygdala will be more likely to exhibit stress behaviors while facing stressors. These findings expand our understanding about the role of amygdala in stress-related behaviors and psychiatric disorders. The world is currently witnessing increasing uncertainty and all kinds of natural disasters are on the rise. Our current findings provide a new venue to identify the vulnerable populations to stress, which has great implications to the society. Timely and precise identification of individuals who are vulnerable to stressors and taking strategies to protect them from the negative impact of stressors is very urgent and important. It helps to reduce waste of public resources, and also helps to decrease the likelihood of potential vulnerable individuals to develop stress-related psychiatric disorders.

## Data Availability

These data are available upon direct request for use by qualified researchers.

## Acknowledgements

This work was supported by the National Natural Science Foundation of China (Nos. 81771473 and 81371476). The authors gratefully acknowledge Karl Friston in the Wellcome Centre for Human Neuroimaging, University College London for his insightful comments on self-connections. The authors also acknowledge Peifu Li, Haixia Mao Jun Chen and the staff in the Department of Psychiatry and Department of Radiology, Renmin Hospital of Wuhan University and Chunlin Yang and the staff in the Department of Radiology, Beijing Anding Hospital for their extensive time and effort in data acquisition.

## Footnotes

1 These data are available upon direct request for use by qualified researchers.

