## supplementary materials for "Amygdala connectivity as a predisposing neural feature of stress-induced behaviour during the COVID-2019 outbreak in Hubei"

**Amygdala connectivity predicts stress-like behavior during the COVID-2019 outbreak**

**^1^Corresponding authors:**

Yuan Zhou

**This file includes:**

Supplementary texts

Table S1

Figure S1 and S2

References

**Supplementary texts**

**I. The development of a scale assessing** **COVID-19 related stress behaviors**

Principal investigators conducted interviews to 10 physicians and students from the Renming Hospital, Wuhan University and Beijing Anding Hospital on the question of “Based on your observation, please list ten most common stress behaviors among general populations during the period of COVID-19”. Principal investigators also collected a total number of 50 news with the key words “COVID-19 related coping behaviors” (in Chinese) on Baidu News (<http://news.baidu.com/>), one of the most prevalent news search engines in China. Content analyses on interview records and news were performed to synthesize the most frequently mentioned COVID-19 related stress behaviors. Investigators merged similar items to form a final pool of behavior sample, which contained fourteen items. Another three independent raters who were psychological students from University of Chinese Academy of Sciences evaluated whether each behavior in the list was (1) a frequently seen and (2) a typical COVID-19 stress behavior among general populations on the Yes/No option. All students rated ‘Yes’ on the high frequency and high typicality for all items.

In the questionnaire, participants indicated the degree to which each of the fourteen items matched their behaviors during the COVID-19 pandemic on a six-point Likert Scale ranging from 1=does not match at all to 6=matches to a great extent (Table S1 in the supplementary materials). The scale had good internal validity with Cronbach’s Alpha amounting to .87 and .89 respectively for the Hubei Cohort and the non-Hubei Cohort. Explorative factor analysis on the combined sample generated one common factor with eigenvalue exceeding 1 and with factor loadings of all items on the common factor exceeding .40.

In order to test the criteria validity of this scale, we also collected the self-report measurements on anxiety and stress during this survey. Trait anxiety and state anxiety were measured by a Chinese version 40-item scale translated and revised from the State-Trait Anxiety Inventory (S-TAI) (1). Perceived stress was measured by a Chinese version 10-item scale translated and adapted from the Perceived Stress Scale-10 (PSS-10) (2). Particularly, participants indicated over the last month the frequency they had each of the negative experience described in each item as a result of COVID-19 pandemic on a five-point Likert scale. The Pearson correlation with the Perceived Stress Scale (PSS) and the State Anxiety Inventory (SAI) amounted to r = .21, p = .024, and r = .25, p = .007 respectively, suggesting the COVID-related stress behaviors scale has good criteria validity. In addition, we used a Chinese version 9-item scale translated from the Patient Health Questionnaire-9 (PHQ-9) (3) to measure depression.

II. **Complementary analysis**

**Participants**

In order to test the validity of the SBSC, we recruited another group of the normal controls in an ongoing fMRI study of major depressive disorders, conducted at Beijing Anding Hospital, Capital Medical University, which began in 2018. Fifty-eight normal controls, who completed MRI scanning before December, 2019, were recruited in the current study. All of these participants resided outside the Hubei province when they were recruited in the original project and half a year before COVID-19 outbreak in Hubei (non-Hubei Cohort).

**Imaging protocol**

MRI scanning was preformed by using a 3.0T Siemens Prisma MRI scanner with a 64-channel phased-array head coil at the radiology department of Beijing Anding Hospital, Capital Medical University. Resting-state functional images were also obtained using EPI sequence (TR = 2000 ms, TE = 30 ms, FA = 90º, matrix = 64 × 64, FOV = 200 × 200 mm^2^, slices number = 33, with a thickness of 3.5 mm, voxel size = 3.13 × 3.13 × 4.2 mm^3^) and 200 volumes were obtained. Structural images were obtained using a magnetization-prepared rapid acquisition gradient echo (MPRAGE) sequence (TR = 2530 ms, TE = 1.85 ms, FA = 15º, matrix = 256 × 256, FOV = 256 × 256 mm^2^, slices number = 192, with a thickness of 1 mm, voxel size = 1 × 1 × 1 mm^3^). During the resting-state scanning for both of the cohorts, all participants were instructed to close their eyes and to focus on nothing in particular.

**Neuroimaging data analyses**

All imaging data preprocessing procedures were the same as the main analyses, except the criteria of “good” volumes of data. That is, we excluded participants who had less than 100 “good” volumes of data (Yan et al. 2013) or whose mean FD was above 3 standard deviations beyond the mean value of the whole sample.

We conducted the functional connectivity analyses as the same procedure as the main analyses. Due to no correlations between the rsFC of the left or right amygdala and the SBSC scores were found (cluster-level FWE *p* < .025), no furthermore effective connectivity was conducted in the non-Hubei Cohort.

Table S1. The scale items of COVID-related stress-like behaviors

| Num. | Items |
| --- | --- |
| 1 | Repeatedly takes temperature. |
| 2 | Frequently washes hands; the frequency or the amount of time on handwashing is higher than past. |
| 3 | Thoroughly disinfects the house everyday |
| 4 | Afraid that the current mask storage is insufficient and the masks lack protective capability. |
| 5 | Wears goggles or other eye protection equipment when going outdoors. |
| 6 | Wears rain coats or other protective clothing when going outdoors. |
| 7 | Often feels tired and uncomfortable. |
| 8 | Rushes to buy or hoards daily necessities and food |
| 9 | Afraid of going outdoors, even if there is a severe food shortage. |
| 10 | Worries that self or family members would be infected. |
| 11 | Gets nervous when thinking about the harm of COVID-19. |
| 12 | Spends considerable amount of time every day reading about news related to COVID-19. |
| 13 | Calls mental support hotlines or seeks for online metal counseling services. |
| 14 | Suspects oneself of being infected by COVID-19. |

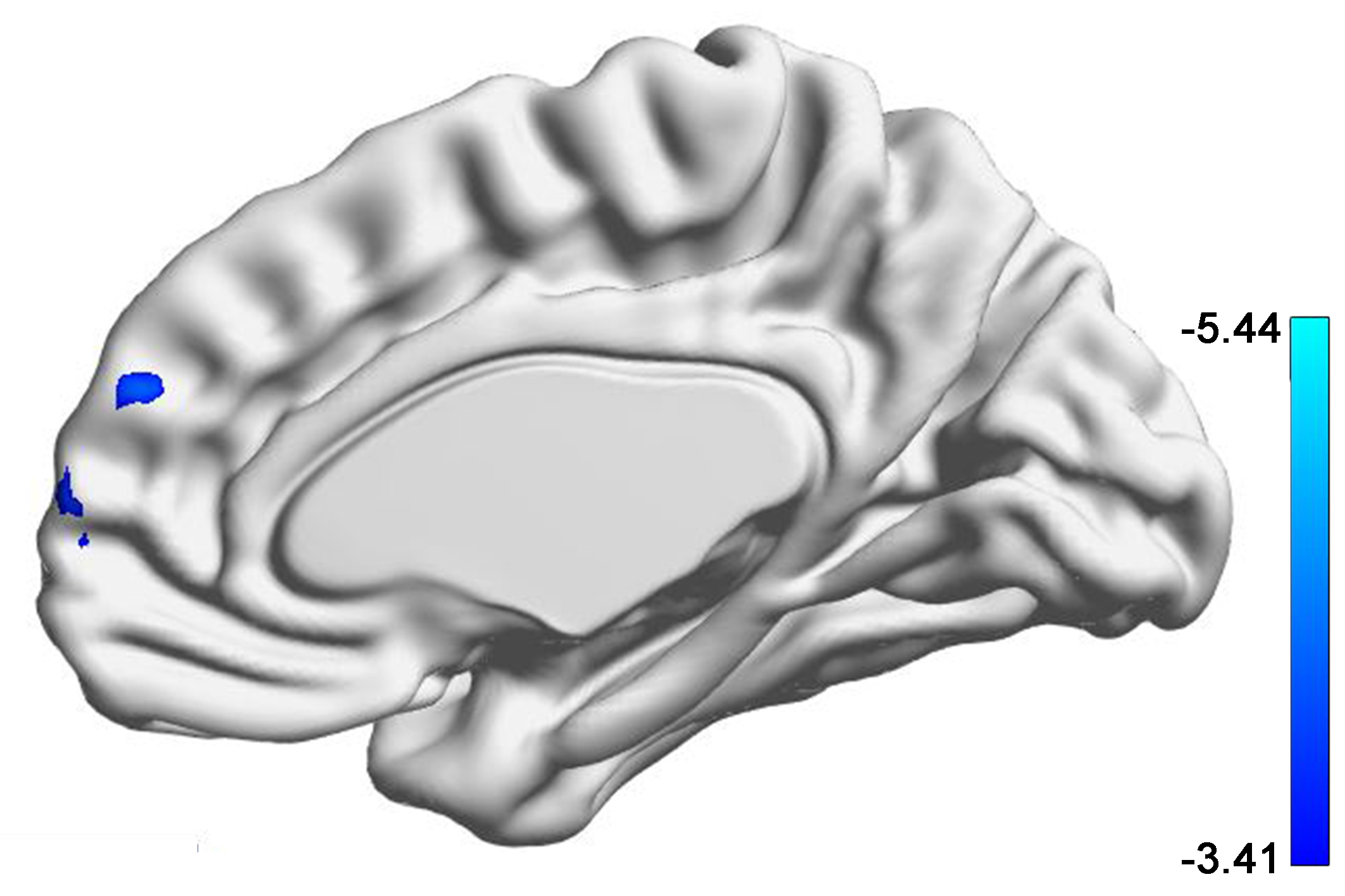

Figure S1. A cluster in the dmPFC whose rsFC with the right amygdala correlated with the stress behaviors during the COVID-19 outbreak at the first survey.

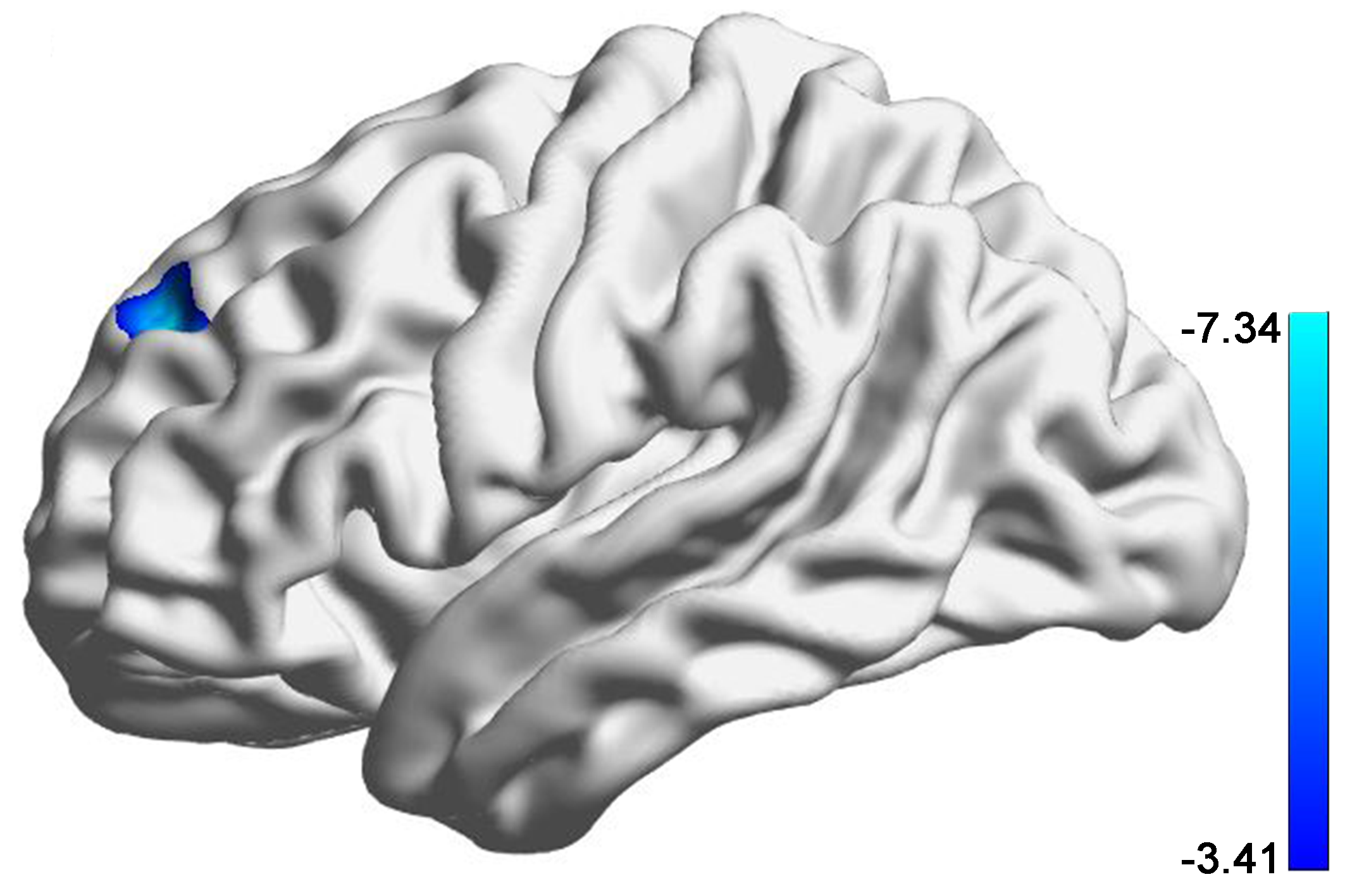

Figure S2. A cluster in the left superior frontal gyrus whose rsFC with the right amygdala correlated with the stress behaviors during the COVID-19 outbreak at the second survey.
